# Lysosomal Storage Dysfunction, a Germline Variants Affecting Pancreatic Ductal Adenocarcinoma Development and Progression

**DOI:** 10.1101/2023.02.15.23286007

**Authors:** Youngil Koh, Hyemin Kim, Seulki Song, Young Hoon Choi, So Young Joo, Hyung Rae Kim, Byul Moon, Jamin Byun, Junshik Hong, Dong-Yeop Shin, Solip Park, Kwang Hyuck Lee, Kyu Taek Lee, Jong Kyun Lee, Daechan Park, Jin-Young Jang, Hyunsook Lee, Jung-Ae Kim, Sung-Soo Yoon, Joo Kyung Park

## Abstract

Lysosome is closely linked to autophagy, which plays a vital role in pancreatic adenocarcinoma (PDAC) tumor biology. This study investigated whether lysosome storage dysfunction (LSD) contributes to PDAC development. Germline putative pathogenic variants (PPVs) in genes involved in lysosome functions were compared between PDAC patients (N=418) and healthy controls (N=845). Furthermore, *Galc*-knockout mouse pancreas organoids and human PDAC organoids were used to evaluate the consequences of PPV status in PDAC development and establishment. LSD PPVs were enriched in PDAC patients (Log2OR=1.65, P=3.08×10^−3^). PPV carriers diagnosed with PDAC were younger than non-carriers (61.5 vs. 65.3 years, P=0.031). Hampered autophagolysosome activity with increased autophagy flux and elevated Ki-67 index were observed following GALC downregulation. RNA sequencing of human PDAC organoids revealed upregulation of metabolism related to LSD. Genetically defined lysosome dysfunction is frequently observed in young-age onset PDACs. Lysosome dysfunction might contribute to PDAC development via altered metabolism and impaired autophagolysosome activity.

## INTRODUCTION

Despite medical advances, pancreatic cancer remains one of the world’s deadliest malignancies. According to Global Cancer Statistics, pancreatic cancer accounts for almost as many deaths (466,000) as cases (496,000) because of its poor prognosis. It is the seventh leading cause of cancer death worldwide.^1^ Important cause for this dismal outcome is delayed diagnosis: only 10-20% of patients with pancreatic cancer are diagnosed with resectable disease.^2^ Accordingly, identifying high-risk populations for an earlier diagnosis and improved survival is necessary. Up to 10% of all pancreatic cancers are estimated to be attributable to inherited risk factors.^3,4^ The genetic basis of familial or hereditary pancreatic cancer can be explained in 21% of families by previously described hereditary cancer genes and in 35% of families by low-frequency variants in other DNA repair genes.^5^ More specifically, germline mutations in genes related to DNA instability, such as CDKN2A, TP53, MLH1, BRCA2, ATM, and BRCA1, are well known to be associated with pancreatic adenocarcinoma (PDAC) development.^6^ Knowledge related to germline variants driven susceptibility is essential for PDAC because it helps define a high-risk population and serves as a rational background for novel anti-cancer drug development. Considering that just around 50% of familial PDAC can be explained with current knowledge, vigorous research is warranted to reveal the underlying genetic mechanism of PDAC further. Besides the above cancer-predisposing genes, lysosomal storage diseases (LSDs) comprise more than 50 disorders caused by mutations in genes involved in the function of endosome–lysosome proteins.^7^ Lysosomes are the main digestive compartments within cells and are closely related to autophagy, a primary intracellular degradation system that derives its degradative abilities from the lysosome.^8^ For cancer cells, lysosome affects growth factor signaling through endocytic degradation of growth factors, their receptors, or signal transduction mediators to modulate signaling output.^9^ Also, defective autophagy is suggested to participate in carcinogenesis, probably due to reduced removal of defective organelles or damaged cells.^10^ Accordingly, there is a chance for a close correlation between lysosome dysfunction and cancer development. Indeed, a previous study showed a possible association between rare variants in LSD genes and cancer.^11^ In the survey, pathogenic variants in LSD genes were significantly enriched in the cancer cohort compared to the average population. Especially, PDAC showed a strong association with germline mutations in several LSD genes, including SGSH, MAN2B1, and IDUA. Therefore, these results coincide with mouse model evidence suggesting that autophagy suppresses cancer initiation.^12^ Also, recently, some chronic diseases of adult onset are known to originate from LSD gene variant heterozygote background. A good example is an association between Parkinson’s disease in GBA mutant heterozygote carriers.^13^ Above all, we hypothesized that some LSD heterozygote carrier status might evoke PDAC via suppressed lysosome dysfunction. In addition, once established, cancer cells use autophagy to promote survival during nutrient stress and recycling of cell components to support a transformed phenotype^14^; PDAC is highly dependent on enhanced lysosome function to facilitate degradation, clearance, and recycling of cellular material delivered by increased rates of vesicle trafficking through autophagy and macropinocytosis.^15-17^ Hence, impaired autophagy activity might contribute to cancer initiation, but the role of altered autophagy function in established cancer cells might work differently. It is well known that autophagy has a biphasic role in cancer initiation and progression.^18^ Bring it all together, the study aimed to investigate the oncogenic effect of LSD heterozygote carrier status in PDAC, 1) evaluate the clinical meaning of LSD gene rare variants in large-scale PDAC and healthy control cohorts, 2) focus on an ethnically homogenous population such as Korean since rare variants analysis is primarily affected by ethnicity, 3) evaluate the functional consequences of LSD gene dysfunction using mouse organoid and human cancer cell lines, 4) assess characteristics of PDAC harboring LSD rare variants to understand the features of LSD-relate PDAC using human PDAC organoids.

## RESULTS

### Baseline characteristics of study patients

The basal characteristics of 418 PDAC patients are presented in Table 1. The age ranged from 35 to 87 years, with a median age of 65. Of those, 243 patients (58.1%) were male, and 175 patients (41.9%) were female. The median value of BMI was 22.7 kg/m2, and 146 patients (34.9%) had diabetes mellitus (DM) at the time of diagnosis. In the laboratory findings, the median value of CEA and CA 19-9 was 2.4 ng/ml and 150.6 U/ml, respectively. According to the 8th edition of the AJCC staging system, 78 (18.7%), 123 (29.4%), 112 (26.8%), and 105 (25.1%) patients were in stages I, II, III, and IV, respectively. Univariate analysis showed that the stage was associated with overall survival, implying that our cohort was a general pancreatic cancer cohort rather than a biased cohort with an unusual disease course.

**Table 1.**
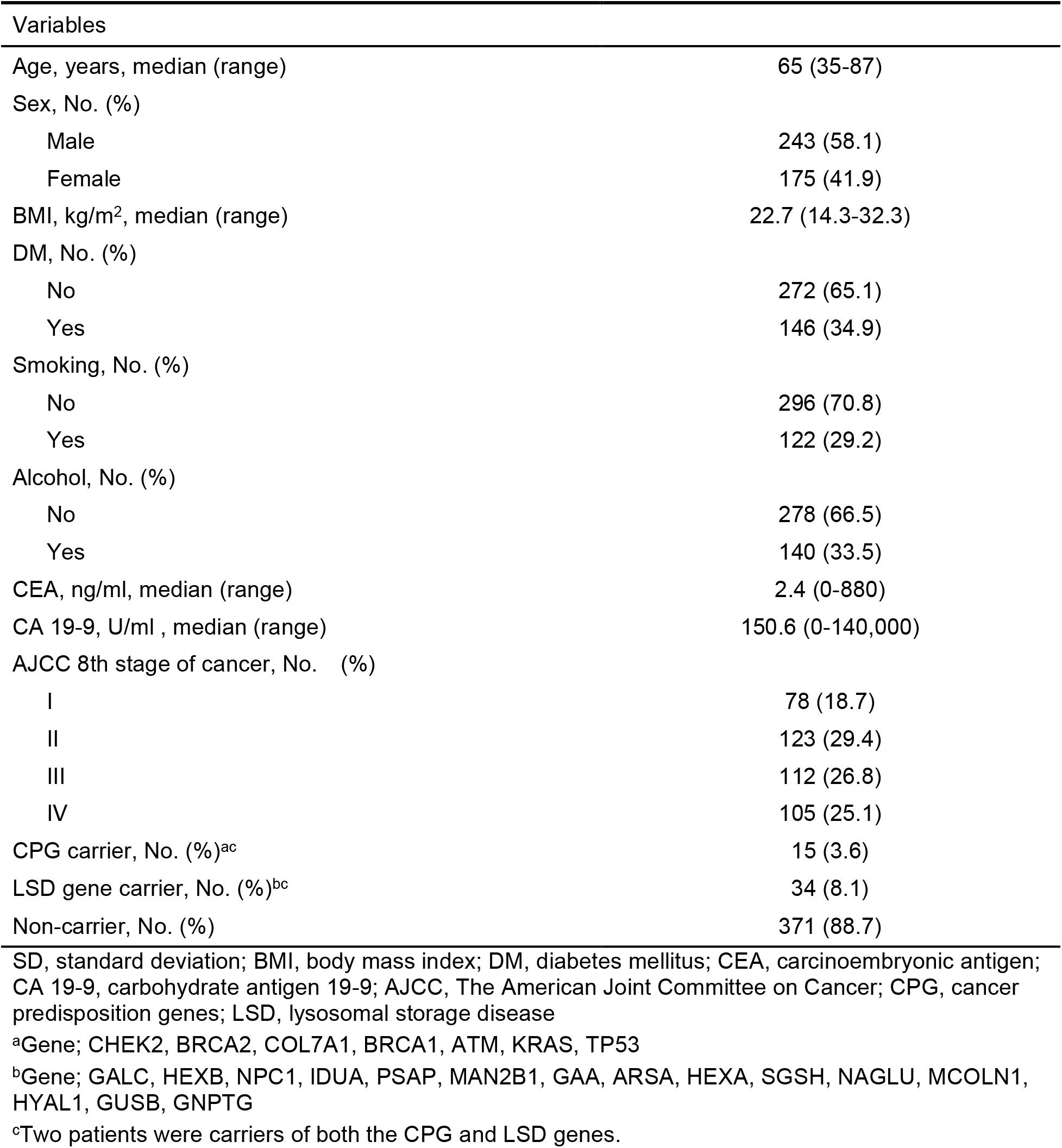
Baseline Characteristics of Patients with Pancreatic Cancer

### Enrichment of LSD germline variants in PDAC cohort

Focusing on genomic loci of 42 LSD genes, 37 rare pathogenic germline single nucleotide variants (SNVs) and insertions and deletions (indels) were detected in 21 out of the 42 LSD genes (*ARSA, GAA, GALC, GLB1, GNPTAB, GNPTG, GUSB, HEXA, HEXB, HGSNAT, HYAL1, IDUA, MAN2B1, MCOLN1, NAGLU, NPC1, PSAP, SGSH, SMPD1, SUMF1*, and *TPP1*). Most variants were protein truncating variants (PTVs; 24 out of 37), and 48.7% were clinically proven pathogenic variants from ClinVar (Pathogenic or Likely Pathogenic; 18 out of 37, Supplementary table 3).

Overall, 34 Korean PDAC patients (8.1%) carried at least one genomic variant, whereas 36 heterozygote carriers (4.3%) were identified among controls history of cancer. The prevalence of cancer with LSD rare heterozygous germline variants remained significant compared to controls (*p* = 6.92 × 10^−3^ by Chi-square test). In particular, the most frequently mutated gene in Korean PDAC patients was *GALC* (3.6%), followed by *HEXB* (1.2%), *GAA* (0.5%), and *NAGLU* (0.5%, Supplementary Table 1). We also estimated the effect of rare germline variants using a regression model after gender and age adjustment using samples with WES data (418 PDAC patients and 352 controls to avoid variant selection bias of technical differences). It revealed PDAC enrichment of LSD variants with a Log2 odds ratio (OR) of 1.65 (*p* = 3.08 × 10^−3^). Of the 418 Korean PDAC patients, 15 (3.6%) had CPG mutations, including *BRCA1/2, ATM*, and *COL7A1*, and two patients had rare pathogenic germline variants on both LSD genes and CPGs (Table 1).

### Early onset of PDAC in patients with LSD gene carriers

We tested whether LSD gene carriers develop PDAC at a younger age than other cancer predisposition genes (CPGs) (Supplementary Figure 1 and Table 2). Two patients who were both CPG and LSD gene carriers were classified as CPG carriers, and the characteristics of each study group were compared. The mean ages at diagnosis of PDAC in CPG carriers and LSD gene carriers were 60.3 and 61.7 years, respectively, significantly lower than 65.3 years in patients without CPG or LSD gene carriers (*p* = 0.025). In addition, for all LSD gene carriers (n = 34), including two who also have CPG mutations, the mean age at diagnosis of PDAC was 61.5 years, which was significantly lower than 65.3 years for non-carriers for either LSD gene or CPG (n = 371) (*p* = 0.031). Other than age, there were no significant differences in clinical characteristics, including tumor stages according to CPG or LSD gene carrier status (Table 2).

**Table 2.** Patient characteristics according to CPG or LSD genes carrier status

| Variable | CPG carrier<sup>a</sup><br>n = 15 | LSD gene carrier<sup>a</sup><br>n = 32 | Non-carrier<br>n = 371 | P-value |
| --- | --- | --- | --- | --- |
| Age, years, mean ± SD | 60.3 ± 7.2 | 61.7 ± 11.5 | 65.3 ± 9.54 | **0.025** |
| Sex |  |  |  |  |
| Male | 9 (2.2) | 22 (5.3) | 212 (50.7) | 0.438 |
| Female | 6 (1.4) | 10 (2.4) | 159 (38.0) |  |
| BMI, kg/m<sup>2</sup>, mean ± SD | 23.0 ± 2.8 | 22.3 ± 2.6 | 22.8 ± 3.0 | 0.628 |
| DM, No. (%) |  |  |  |  |
| Yes | 5 (1.2) | 14 (3.3) | 127 (30.4) | 0.551 |
| No | 10 (2.4) | 18 (4.3) | 244 (58.4) |  |
| Smoking, No. (%) |  |  |  | 0.597 |
| Yes | 3 (0.7) | 11 (2.6) | 108 (25.8) |  |
| No | 12 (2.9) | 21 (5.0) | 263 (62.9) |  |
| Alcohol, No. (%) |  |  |  | 0.809 |
| Yes | 4 (1.0) | 10 (2.4) | 126 (30.1) |  |
| No | 11 (2.6) | 22 (5.3) | 245 (58.6) |  |
| CEA, ng/ml, mean ± SD | 9.7 ± 25.5 | 4.0 ± 6.5 | 9.9 ± 55.2 | 0.872 |
| CA 19-9, U/ml, mean ± SD | 555.0 ± 1136.3 | 4092.3 ± 8559.6 | 2677.3 ± 11499.8 | 0.588 |
| AJCC 8th stage of cancer, No. (%) |  |  |  | 0.840 |
| I | 4 (1.0) | 8 (1.9) | 66 (15.8) |  |
| II | 4 (1.0) | 11 (2.6) | 108 (25.8) |  |
| III | 4 (1.0) | 7 (1.7) | 101 (24.2) |  |
| IV | 3 (0.7) | 6 (1.4) | 96 (23.0) |  |

CPG, cancer predisposition genes; LSD, lysosomal storage disease; SD, standard deviation; BMI, body mass index; DM, diabetes mellitus, CEA, carcinoembryonic antigen; CA 19-9, carbohydrate antigen 19-9; AJCC, The American Joint Committee on Cancer
<sup>a</sup>Two patients who were carriers of both the CPG and LSD genes were classified as CPG carriers.

### Increased autophagy flux with GALC gene variants in PDAC cell lines

The endogenous activity of *GALC* in PK59 (*GALC* mutation), HPAFII (*GALC* wild-type), and Capan-1 (*GALC* wild-type) cells was examined by lysosomal GALC enzyme activity analysis, showing low activity of *GALC* in PK59 cells (Figure 1A). Also, the expression of LC3B-II, the most widely used autophagosome marker, was examined by immunoblotting after treatment of chloroquine (CQ), an autophagic inhibitor, to monitor autophagy flux. The enhanced amount of LC3B-II by CQ was prominent in PK59 cells (Figure 1B and 1C). Moreover, we constructed PK59 and HPAFII cells stably expressing GFP-LC3 and monitored the number of LC3 after CQ treatment. The relative increase of GFP-expressed cells was more remarkable in PK59 cells than in HPAFII cells (Figures 1D and 1E). These results indicate the enhanced autophagy flux in *GALC*-mutated PDAC cells compared to *GALC* wild-type PDAC cells.

**Figure 1.**
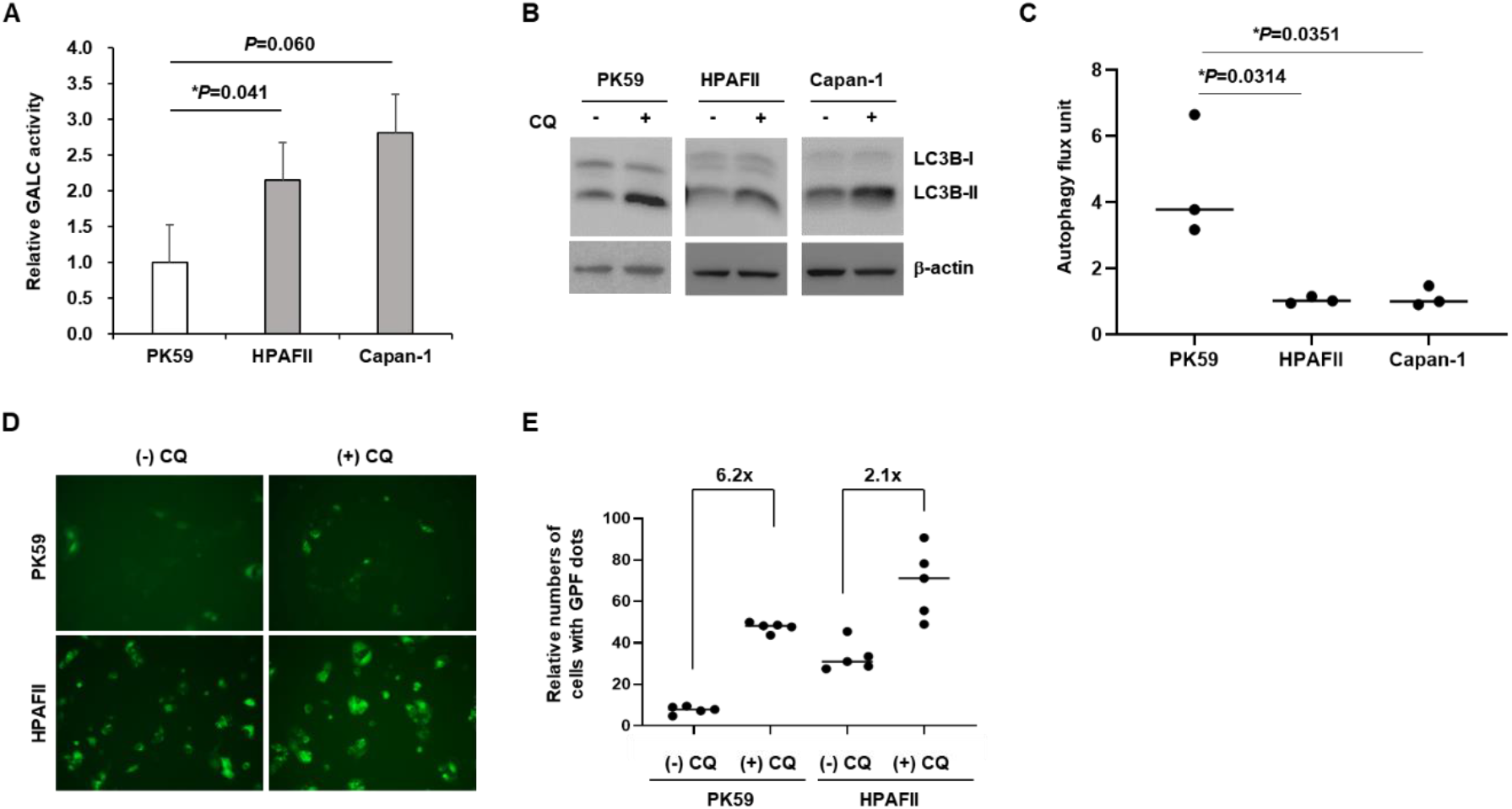
Enhanced autophagic flux in GALC-defective pancreatic ductal adenocarcinoma (PDAC) cells. (A) Endogenous activity of GALC in multiple PDAC cell lines was measured by lysosomal GALC analysis. (B) PK59, HPAF-II and Capan-1 cells were treated with or without chloroquine (CQ) for 1 hour to monitor autophagy flux. Expression levels of LC3B in PDAC cell lines was detected in three independent immunoblottings. (C) Autophagy flux unit represents a buildup of LC3B-II according to CQ treatment. Densitometric analysis was performed to measure the band intensity of LC3B using ImageJ. (D) PK59 and HPAF-II that stable expressed GFP-LC3 cells were treated with CQ for 5 hours and GFP-LC3 dots were measured with a fluorescence microscopy. (E) The numbers of GFP-LC3 dots were counted manually from the fluorescent images. The indicated numbers above graphs described an increased ratio of LC3 dots in CQ treatment condition relative to no-treatment.

### Defective lysosomal function in *Kras*^*G12D*^*/Galc* knockout mouse organoids

To investigate the effect of *GALC* variants on autophagy regulation and lysosomal function in PDAC, pancreatic organoids from *KrasG12D* mice were cultured, and *Galc* was eliminated by CRISPR/CRISPR-associated protein 9 (Cas9) to create pancreatic *KrasG12D/Galc* knockout (KO) organoids (Supplementary Figure 2). The expression of LC3B-II was slightly increased in *KrasG12D* organoids, which was markedly augmented in *KrasG12D/Galc* KO organoids accompanied by decreased p62 expression and reduced mTOR phosphorylation (Figure 2A). In addition to detecting autophagic flux, lysosomal degradation was examined. The lysosomal dysfunction was indicated as LAMP1, a lysosome marker, was remarkable in *KrasG12D/Galc* KO organoids (^**^P<0.01, Figure 2B and 2C), and it led to ubiquitinated proteins accumulation (Figure 2F). Furthermore, the number of autophagosomes was higher in *KrasG12D/Galc* KO organoids than in wild-type or *KrasG12D* organoids, and the number of autolysosomes was not different among the three organoids (Figures 2E and 2F). Thus, defective *GALC* expression enhanced autophagic flux and autophagosome formation. Still, autolysosome formation and degradation were not followed due to the lower lysosome and lysosomal dysfunction, causing an accumulation of ubiquitinated proteins in *KrasG12D/Galc* KO organoids.

**Figure 2.**
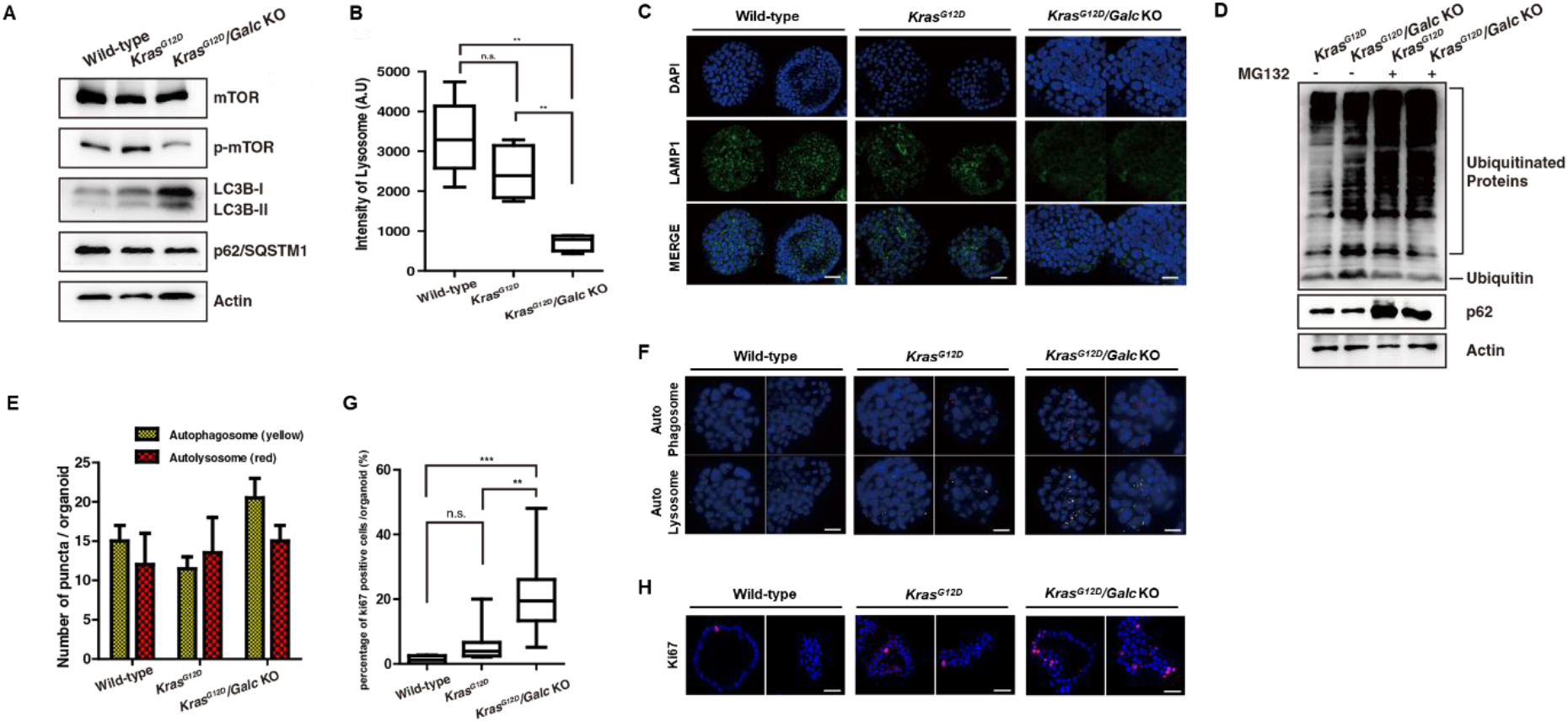
Defective lysosomal function and increased proliferation in KrasG12D/Galc knockout mouse pancreatic organoids. (A) The expression of mTOR, phosphorylated mTOR (p-mTOR), LC3BI/II and p62 for autophagic regulation was examined by immunoblotting. (B and C) The intensity LAMP1, a lysosomal marker, was measured with immunofluorescent staining. p^**^<0.01. (D) Proteasomal degradation levels were detected after MG132, a proteasome inhibitor, treatment. (E and F) Fluorescence-labeled autophagosomes and autolysosomes were detected in mouse pancreatic organoids with KrasG12D or Galc knockout. p^**^<0.01, p^***^<0.001. (G and H) The number of Ki67, a proliferation marker, was counted with confocal fluorescence microscopy. p^**^<0.01, p^***^<0.001.

### Increased proliferation of *Kras*^*G12D*^*/Galc* knockout mouse organoids

To understand the regulatory mechanism of lysosomal dysfunction affecting pancreatic tumorigenesis, we observed the gene expression pattern of organoid samples derived from *KrasG12D and KrasG12D/Galc* KO mice using RNA-seq data. The expression of GALC was decreased in *KrasG12D/Galc* KO mouse as expected (median expression value of *KrasG12D* and *KrasG12D/Galc* KO mouse: 1.04 vs. 5.13, *p* = 0.05 by Wilcoxon rank sum test, Supplementary Figure 3A). In differentially expressed genes analysis using RNA-seq data, the majority (81.5%) of genes, including the *MIRG, PIGR, SYT14, PLEKHD1*, and *SLC2A1*, were down-regulated in *KrasG12D/Galc* KO organoids. In addition, low gene expression of Monoamine oxidase A (*MAOA*), a key factor regulating apoptosis/autophagy by targeting the repressor element-1 silencing transcription factor (*REST1*), was also identified in *KrasG12D/Galc* KO mouse organoids. On the other hand, the expression of genes including *STAG3, GATA5*, and *CACNB2*, which are associated with cell proliferation or tumorigenesis, and *PRAP1*, which is primarily involved in the apoptosis inhibition of tumor cells, was increased in *KrasG12D/Galc* KO mouse organoids (Supplementary Figure 3B).

Regarding cell proliferation, we tested the Ki67 expression in pancreatic mouse organoids. As a result, Ki67-positive proliferating cells were considerably increased in *KrasG12D/Galc* KO organoids, and they were much lower in wild-type and *KrasG12D* organoids (Figure 2G and 2H, Supplementary Figure 4). Furthermore, gene set enrichment analyses (GSEA) with MSigDB Gene Ontology revealed that *KrasG12D/Galc* KO mouse organoids were strongly associated with gene ontology (GO) terms such as *RAS*-protein signaling, cytokine production, and cell death (Supplementary Figure 3C). However, the regulation of translation, RNA processing, protein ubiquitination, and glycolysis were significantly down-regulated in the *KrasG12D/Galc* KO organoids.

### Evaluation of pathways associated with LSD germline variants in human PDAC

Finally, to investigate the functional consequences of lysosomal dysfunction in established PDAC, we conducted RNA-seq from LSD PTV carriers (N = 2, both were *GALC* heterozygote carriers) and non-carriers (N = 27) using our patient-derived PDAC organoids. Differential expression analysis identified 240 differentially expressed genes (DEGs), of which 68 were up-regulated, including *S100P, AFAP1-AS1, TFCP2L1*, and *RASA3*, and 172 were downregulated, including *APOM, APOC1, ASB4, TF*, and *PLG* in LSD PTV carrier PDAC organoids (log2FoldChange > 1, false discovery rate (FDR) < 0.05) (Figure. 3A). Interestingly, a GSEA showed that LSD PTV carrier PDAC organoids were strongly up-regulated in amino acid metabolic biosynthesis-related pathways including primary bile acid biosynthesis, steroid hormone biosynthesis, linoleic acid metabolism, alanine, aspartate and glutamate metabolism and glycerolipid metabolism in KEGG (Figure. 3B). Furthermore, we could also identify the DNA repair system (nucleotide excision repair, mismatch repair, and homologous recombination) up-regulation in LSD PTV carrier PDAC organoids. In addition, metabolic up-regulation was confirmed through the GO method from a biological point of view that occurs within cells in LSD PTV carrier PDAC organoids (Figure. 3C). However, most cell-to-cell signaling pathway and organ development pathways were down-regulated in LSD PTV carrier PDAC organoids (Figure. 3C). Moreover, we conducted drug test with patient-derived PDAC organoids (one *GALC* carrier, two non-LSD carriers) to examine the response to treatments (Figure 3D). PDAC patient-derived organoids with *GALC* carrier tend to be more resistant to drugs, including gemcitabine, nab-paclitaxel (Abraxane), paclitaxel, irinotecan, liposomal irinotecan (Onivyde), and Olaparib compared to non-LSD carriers. In addition, human PDAC cell lines with LSD carrier such as *GALC* carrier (PK59) was more resistant to drugs, including gemcitabine, nab-paclitaxel (Abraxane), irinotecan, liposomal irinotecan (Onivyde), oxaliplatin, and Olaparib (Supplementary Figure 5).

**Figure 3.**
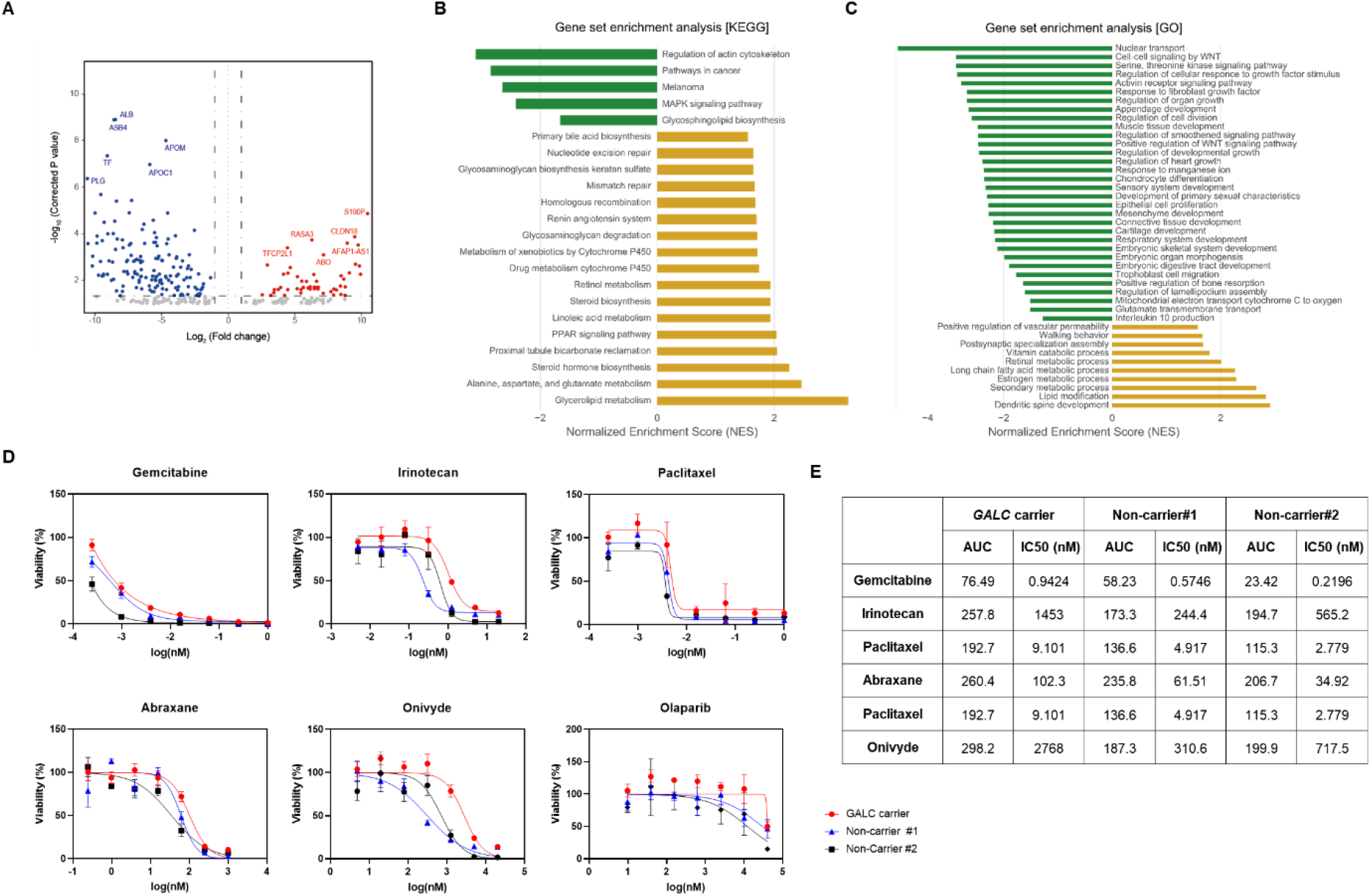
Evaluation of GALC carrier in human pancreatic ductal adenocarcinoma (PDAC) organoids. (A) Volcano plot with RNA sequencing showed differentially expressed genes between GALC carrier PDAC organoids (n=2) and non-carrier PDAC organoids (n=27). (B and C) The biological process, cellular components and molecular function were identified from the GO and KEGG enrichment analysis. GO, Gene Ontology; KEGG, Kyoto Encyclopedia of Genes and Genomes. (D) Therapeutic drugs were treated to human PDAC organoids for 7 days, and cell viability was accessed using an adenosine triphosphate monitoring (triplicates). Dose– response curves (DRCs) were fitted, and the area under the curve (AUC) for each DRC and the half-maximal inhibitory concentration (IC50) values (nM) were calculated.

## DISCUSSION

Germline mutations in genes confer a risk for cancer development and cause early onset of cancer.^19^ So far, most genes involved in carcinogenesis have functions related to cell division and proliferation. Here, we investigated the oncogenic effects of LSD heterozygosity in PDAC patients: 1) our study evaluated the clinical meaning of LSD gene rare variants in large-scale PDAC patients and healthy control cohorts, 2) the functional analyses were conducted using mouse pancreas organoids, LSD carrier PDAC cell lines, and patient-derived PDAC organoids. First, LSD PTV carriers were enriched in the PDAC population compared to healthy control and developed PDAC at a younger age than non-carrier PDACs. Second, we showed paradoxically increased autophagy flux due to impaired autophagolysosome activity which might contribute to PDAC development using mouse organoids and PDAC cell line experiments. Third, LSD PTV carrier patient-derived PDAC organoids showed upregulation of metabolic pathways, which supports that genes involved in lysosomes might also contribute to PDAC development via altered autophagy activity.

The enrichment of germline LSD PTV was around three times (Log2Odds Ratio 1.6-1.8) in the Korean PDAC population compared to healthy controls. This enrichment is also accompanied by the younger age of onset of PDAC by 3.6 years compared to non-LSD PTV carriers like other CPGs. A previous study reported the enrichment of six representative germline PTVs, including BRCA1/2 and TP53, in the PDAC population compared to the general population, with ORs ranging from 2.58–12.3 (all ORs were more significant than five except for 2.58 for BRCA1)^6^ implying that the oncogenic potential of germline LSD PTV is weaker than the conventional CPG’s. The dysfunction in lysosome and autophagy is an emerging hallmark of cancer, unlocking phenotypic plasticity^20^ but not a classic hallmark of cancer – i.e., cell proliferation and replication.

Technically, because of the genetic diversity of rare variants for diseases,^21,22^ it is crucial to address ethnic factors in this kind of analysis. We eliminated this issue by focusing on the single ethnic Korean population for germline PTV analysis. Korean is entirely genetically homogeneous, and comparing the PTV frequencies across cohorts is feasible. In PTV analysis, *GALC, HEXB, GAA*, and *NAGLU* were the genes with high frequency in the Korean PDAC population. Of these, the contribution of *GALC* PTVs (rs138577661 and rs200607029) was considerable in our study, and indeed these two are well-known variants for Krabbe’s disease in East Asian populations.^23^ The functionality of the rs138577661 variant was also confirmed by enzymatic testing of the PDAC cell line (PK59) harboring the exact variant. All in all, variant-level data also suggests the importance of ethnicity in this rare PTV study. Hence, extrapolating our data to other ethnic populations should be done cautiously.

Among various cancers, PDAC has been selected as a cancer type for the experiment in this study based on a previous study revealing the possible link between LSD PTV and PDAC^11^ and its well-known biology related to the lysosome, autophagy,^24^ and abnormal metabolism.^25^ For tumor initiation and development, dysfunctional lysosome impedes autophagolysosome activity of abnormal cells resulting in cancer cell survival and growth.^26^ From the viewpoint of cell death, either lysosomal or autophagic cell death is not adequately working for cancerous cells.^27^ Indeed, much evidence suggests that dysfunctional lysosome contributes to cancer initiation.^14,28^ In this sense, our result generates substantial evidence that *GALC* knock-down promotes cell proliferation (elevated Ki-67) in cooperation with *KRAS* mutation in the pancreas organoid. Also, we observed that the function of autophagolysosome is lowered, which subsequently increases autophagy flux as feedback, suggesting an altered autophagy activity with *GALC* downregulation. Considering lysosome’s cellular function, we hypothesize that cooperation with oncogenic mutations such as *RAS* and lysosomal dysfunction is necessary for carcinogenesis. However, as we did not observe full features of cancer in mouse organoid, future studies using the knock-down mouse of the lysosome-related gene is necessary to confirm spontaneous cancer development.

On the other hand, once the tumor is established, autophagy activity is vital for maintaining cancer in many ways. Especially for PDAC, the nutrient-poor and hypoxic condition makes cancer cells thrive in the harsh environment, which in turn makes autophagy function important to recycle nutrients.^25^ Also, in PDAC, a well-known process called macropinocytosis plays a similar role to autophagy. Both autophagy and macropinocytosis rely on the function of lysosomes for the final degradation of products. It is recently known that macropinocytosis is induced by the blockade of autophagy activity in PDAC via *NRF2* triggering,^29,30^ suggesting their compensatory roles. We interpret that the increased autophagy flux in PDAC originates from lysosomal dysfunction to compensate for hampered autophagolysosome. This increased autophagy flux might play a favorable position in established cancer cell survival with paradoxically increased autophagy and/or macropinocytosis function in the cancer cell. Indeed, RNA-seq analysis revealed that several metabolic pathways were up-regulated in organoids from PDAC patients with LSD PTV carriers, suggesting the enhanced metabolic utility in the established PDAC with lysosome dysfunction. We believe further studies are warranted to generate direct evidence for our interpretation.

Moreover, LSD PTV carriers tended to be less sensitive to drugs in PDAC cell lines and human PDAC organoids. The expression of S100P is known to be associated with drug resistance, metastasis, and poor clinical outcomes.^31^ S100 calcium-binding protein P (S100P) promotes pancreatic cancer growth, survival, and invasion, and its intracellular level affects the resistance against 5-fluorouracil treatment in vitro.^32^ Also, high expression of long non-coding RNA, actin filament associated protein 1 antisense RNA1 (*AFAP1-AS1*) is associated with poor survival and short-term recurrence in PDAC. Knock-down of *AFAP1-AS1* attenuated PDAC cell proliferation, migration, and invasion.^33^ Apolipoprotein M (APOM) can suppress the proliferation and invasion of hepatocellular carcinoma,^34^ breast cancer^35,^ and larynx carcinoma.^36^ These coincided with up-regulated *S100P* and *AFAP1-AS1* and down-regulation of APOM in RNA-seq data of LSD PTV carrier organoids.

From a genetics perspective, our result implies that the heterozygote status of LSD genes is related to cancer development. Accordingly, we checked whether LSD genes follow Knudson’s two-hit hypothesis,^37^ like most CPGs. In copy number analysis using matched tumor sequencing data, we did not observe a significant loss-of heterozygosity in PDAC patients with LSD PTV’s (data not shown). It can be considered that the biphasic role of lysosome and autophagy in PDAC initiation and progression from our result. Loss of whole autophagy function will not be suitable for cancer cell survival in the PDAC microenvironment. Also, it should be mentioned that we are reporting a piece of parallel evidence that LSD PTV carriers can develop an adult-onset chronic disease such as PDAC other than classic LSD phenotype, as already known in Parkinson’s disease development of GBA rare variant heterozygote carriers.^13^

Lastly, many attempts to develop therapeutics targeting autophagy or macropinocytosis are ongoing in PDAC. In addition, nano-drugs are actively investigated in PDAC. Because all of these modalities are closely related to lysosomal function, our result can serve as a cornerstone for novel therapeutics in PDAC. There is also a chance that lysosomal dysfunction in PDAC might serve as a biomarker for these treatments. Lysosomal dysfunction in PDAC might serve as a biomarker for these treatments

Here, we would like to address that genetically defined lysosome dysfunction is frequently observed in young-age onset PDACs. Lysosome dysfunction might contribute to PDAC development via impaired autophagolysosome activity. In established PDAC, lysosomal dysfunction is closely related to increased autophagy flux and up-regulated metabolism.

## MATERIALS AND METHODS

### Study cohort

Four hundred eighteen patients with PDAC diagnosed from November 2011 to August 2020 at Samsung Medical Center (SMC, n=222) and Seoul National University Hospital (SNUH, n=196) were prospectively enrolled and followed up until the end of 2021. Clinical and laboratory information was collected from electronic health records (EHR). We have prospectively built the cancer-free normal control cohort (CFNC) from 845 healthy volunteers in the SNUH healthcare checkup center. The age of all healthy controls was over 50 without cancer history proven by EHR. This study was conducted under the principles of the Declaration of Helsinki, and the study protocol was approved by the institutional review board (IRB) of SMC and SNUH. All patients and volunteers provided written informed consent, and all specimens were collected according to IRB regulations and approval (IRB No. 2018-12-065, 1705-031-852).

### DNA and RNA sequencing

Genomic DNA (gDNA) collected from peripheral blood was extracted using a QIAamp DNA tissue kit (Qiagen, USA), according to the manufacturer’s recommendations. To evaluate the status of LSD germline variants in study cohorts, either targeted panel or whole exome sequencing (WES) was used. In CFNC cohorts, 493 patients underwent targeted LSD panel sequencing instead of WES to identify LSD germline variants. For WES, DNA from 352 CFNC subjects and 418 PDAC patients was sequenced using the Illumina NovaSeq6000 sequencer (Illumina, San Diego, CA). Library capture was conducted with an IDT probe. For LSD panel sequencing, the DNA of 493 subjects from the CFNC cohort was sequenced using the Illumina Hiseq2500 platform. Libraries were constructed using an ACCEL-NGS 2S DNA library kit from Swift Biosciences, including 42 LSD genes (Supplementary Table 1). For RNA sequencing, total RNA was isolated using RNeasy Mini Kit (Qiagen, USA). TruSeq Stranded mRNA (Illumina, San Diego, CA) was used to prepare RNA-sequencing libraries. The 150 bp paired-end sequencing of these libraries was progressed using NovaSeq 6000 Sequencing System (Illumina, San Diego, CA). The quality of these cDNA libraries was evaluated with the Agilent 2100 BioAnalyzer (Agilent, CA, USA). Quality confirmation was performed using NGS QC Toolkit (version 2.3.3).

### SNVs and INDEL calling

Sequence reads were aligned to the human reference genome (hg19) using the Burrows-Wheeler Aligner-MEM v0.7.10 algorithms.^38^ Conversion to BAM was using Picard v1.130. For indel realignment, duplicated fragment elimination and base quality score recalibration was assessed using the Genome Analysis Tool Kit (GATK v3.8.1, Broad Institute).^39^ Using the CollectHsMetrics of Picard tool, filtering of low-quality reads was performed. Single nucleotide variants (SNVs) and insertions and deletions (indels) were detected using HaplotypeCaller of GATK v3.8.1. Variants in each sample’s genomic variant calling format (GVCF) were merged, and a joint calling approach was also performed with GATK. Finally, sequencing errors were filtered out by assessing the variant quality score recalibration (VQSR), GATK’s statistical modeling approach for variant filtration.

### Calling of putative pathogenic germline variants

To generate a consistent set of variant filtration and functional annotation, we annotated the compiled variants in the VCF file using ANNOVAR^40^ to perform filter-based functional annotation and Variant Effect Predictor (VEP)^41^ to explicate the gene-based information of canonical transcripts. In addition, to ensure rare germline variant only, we performed variant filtration with allele frequency (AF) information from The Genome Aggregation Database (gnomAD)^42^ (gnomad.exomes.r2.1.1). We have extracted the potentially damaging variant by substituting two conditions: (1) Tier1 variants were defined as protein truncating variants (PTVs), including splice donor and acceptor sites, frameshift indels, stop gain and lost variants, as well as not Benign or Likely benign loci annotated with Clinvar.^43^ (2) Genetic variants with well-known clinical risks (pathogenic, Likely pathogenic, association, and risk factor) and related phenotypes clearly defined in Clinvar as Tier 2.

### Differentially expressed gene analysis using transcriptomic data

Whole transcriptome sequencing (WTS) reads of pancreatic tumor organoids were aligned to the human reference genome (hg19) using spliced transcripts alignment to a reference (STAR) version 2.5.3a.^44^ Expression count was estimated using the RNA-seq by Expectation Maximization (RSEM-1.3.0)^45^ and normalized using the EdgeR TMM method before running differential expression analysis. Estimation of differential expression between LSD gene variant carrier and non-carrier was performed using gene-specific read counts with an R package platform called DESeq2.^46^ The log2 scale variance stabilizing transformation was completed with differentially expressed genes (DEGs). Gene set enrichment analysis (GSEA) was performed using the java GSEA application version 3.0. Functionally meaningful pathways were explored using the KEGG^47^ and gene ontology (GO) database of MSigDB^48^ resources.

### Copy number alteration analysis

We determined somatic copy-number alterations (CNA) across 42 LSD genes using CNVkit (https://github.com/etal/cnvkit) by comparing the tumor organoid DNA bam files to the germline samples (matched normal pancreatic blood). Mean read depths for each target (interval) were computed and normalized against the single reference of pooled standard samples, then calculating the B-allele frequency. The observed log2 copy number ratios of the region < -0.4 were derived as copy number loss, and CAN segments were visualized using the copy number package.

### Cell lines

We screened for rare pathogenic germline variants in 42 LSD genes among PDAC cell lines from the Cancer Cell Line Encyclopedia (CCLE) database.^49^ PK59 cell line with a GALC mutation (rs138577661, rs137854543) was maintained in RPMI1640 medium with 10% FBS. HPAFII and Capan-1 cell lines with wild-type GALC were maintained in Eagle’s Minimum Essential Medium with 10% FBS and Iscove’s Modified Dulbecco’s Medium with 20% FBS, respectively. All media and FBS were purchased from Gibco.

### Galactocerebrosidase (GALC) enzyme activity analysis

According to the manufacturer’s instructions, endogenous GALC activity was detected using a Lysosomal Galactocerebrosidase Analysis kit (M2774, MarkerGene). Briefly, 4×10^6^ cells were harvested with provided buffer and lysed using 2 × 30 sec— sonication cycles from the BioruptorTM Pico. First, the lipidic fluorogenic substrate in the kit was incubated with 50ug of protein for 2 hours at 37°C. Then, the fluorescence signal at Ex/Em=365/454 nm was detected by a fluorometer (SYNERGY/HTS, BioTek). The value of fluorescence from PK59 cells normalized the relative GALC activity.

### Autophagic flux analysis

To detect the LC3B signal, 3∼4×10^5^ cells were seeded into 6-well plates. The next day, cells were washed twice with pre-warmed PBS before treatment of 100μM CQ (C6628, Sigma) for 1 hour. LC3B (L7543, Sigma) and β-actin (sc-47778, Santa Cruz) were detected by immunoblotting. Chemiluminescent signals were detected and visualized by LAS3000 (Luminescent Image analyzer, FUJIFILM). Autophagy flux unit (A.F.U. = [LC3B-II/LC3B-I]CO(+)/[LC3B-II/LC3B-I]CO(-)) was calculated by analyzing band intensities of LC3B-I and LC3B-II from each three independent experiments using ImageJ software.

PK59 and HPAF-II cells stalely expressing GFP-LC3 were constructed with retrovirus infection (pBABEpuroGFP-LC3, 22405, Addgene). After treatment with CQ (100μM) for 5 hours, GFP signals from LC3 were analyzed by a fluorescence microscope (Inverted Microscope Eclipse Ti-S, Nikon). Five views of images were selected to count the numbers of total and GFP-LC3 positive cells manually.

### Mouse pancreas organoids culture

Pancreatic ducts were isolated from Wild-type and Kras^G12D^ mice and lysed by collagenase P (Roche) and DNase I (Worthington Biochemical Corp.). Lysed pancreatic ductal cells were seeded in a matrigel matrix (Corning) and grown in a culture medium. The culture medium is based on Advanced DMEM/F12, B27TM Supplement (Gibco), GlutaMAXTM (Gibco), Rspo1, mEGF (Peprotech), mNoggin (Peprotech), hFGF10 (Peprotech), N-acetylcysteine (Sigma-Aldrich), A83-01 (Tocris) and nicotinamide (Sigma-Aldrich). To generate Galc knockout mouse pancreas organoids, lentiviral CRISPR/Cas9 was used. sgRNA targeting *Galc* was cloned into the lentiCRISPRv2 vector. LentiCRISPRv2-sgGalc, pMD2G, and psPAX2 were transfected into 293FT. The lentiviruses were collected from cells and concentrated with a Lenti-XTM concentrator (Takara). Lentiviruses were transduced to organoids and dissociated into single cells by TrypLE (Gibco). The transduced cells were recovered in a matrigel matrix and selected with two ug/ul of puromycin. The generation of Galc Knock out mouse pancreas organoids was confirmed by Reverse transcription polymerase chain reaction (RT-PCR) using the following primers: Galc_F: 5’-AGG TCT CCA GCG AGT GAG AAT CAT AG-3’, Galc_R: 5’-TGT GTG AGC TGA TAC CCA GAT AGG AG-3’.

### Immunostaining and In-Situ hybridization (ISH)

Organoids were isolated from matrigel using Cell Recovery Solution (Corning) and washed with cold PBS. The isolated organoids were fixed in 4% PFA for 1 hr, permeabilized in 1% PBS-T (Triton X-100) for 1 hr, and blocked for 1 hr at RT in blocking solution (3% BSA in 0.2% PBS-T). Primary and secondary antibodies (Supplementary Table 2) were incubated at 4°C overnight. The organoids were mounted with VECTASHIELD Antifade mounting solution with DAPI (vector) and imaged using Zeiss LSM 700 confocal microscope. The images were processed with Image J. The fixed organoids were embedded in a paraffin block and subjected to immunohistochemistry (IHC) staining. IHC staining was performed using OptiView DAB IHC Detection Kit (Ventana) as manufacturer’s instruction. Ki67 antibody (MA5-14520, Invitrogen) was used for IHC, and a mouse Galc probe (NM_008079.4) was used for ISH.

### Human organoid culture

PDAC specimens were homogenized with GentleMACSTM tissue dissociator using a human tumor dissociation kit (Miltenyi Biotec) according to the manufacturer’s instruction, which was filtered with a 70 μm strainer. Suspended cells were plated with Matrigel (Corning) and grown in complete medium: advanced DMEM/F12 supplemented with GlutaMAXTM, containing 10mM HEPES, Antibiotic Antimycotics, B27 supplement, N2 supplement (all from ThermoFisher), 1mM N-acetylcysteine, 10mM Nicotinamide (all from Sigma-Aldrich), 60 ng/ml murine Wnt-3a, 500 ng/ml human R-spondin 1, 10nM human Gastrin, 50ng/ml human Noggin, 50ng/ml human epidermal growth factor (EGF), 100 ng/ml, human fibroblast growth factor 10 (FGF10), 0.5 μM A83-01 (all from Peprotech), 1x Primocin (InvivoGen) and ten μM Y-27632.

Organoids were cultured at 37 °C humidified incubator, and the culture medium was partially changed twice a week. PDAC organoids were seeded into 384-well plates (500 cells/well) with technical duplicates and treated with therapeutic drugs for seven days. Cell viability was accessed using an adenosine triphosphate monitoring system based on firefly luciferase (ATPlite 1step; PerkinElmer) and estimated by EnVision Multilabel Reader (PerkinElmer). The relative cell viability for each dose was obtained by normalization with dimethyl sulfoxide (DMSO) per plate.

### Statistical analysis

This research used the linear regression model to assess the association between PTVs in each gene and phenotypic characteristics.

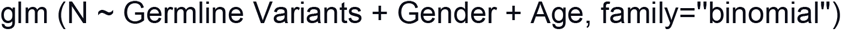

Where: N = case (1) or control (0), Germline Variants = indicating the number of samples that carries rare pathogenic germline variants, Gender = Male (0) and Female (1), and the patients’ age of diagnosis was used as an input for the regression model. A chi-square test was performed for most of the prevalence comparisons. Still, the logistic regression model was used to determine the risk of rare genetic variants in cancer patients compared to the normal control with a significance cutoff of p<0.05. The chi-square and Fisher’s exact tests evaluated the association between categorical variables. The independent t-test and one-way analysis of variance were used to assess the association between continuous variables. Survival analysis was carried out using the Kaplan–Meier method. All statistical analyses were done using either SPSS version 25.0 (IBM SPSS Statistics, Armonk, NY: IBM Corp) or the R language environment (http://www.r-project.org).

## Data Availability

All data produced in the present study are available upon reasonable request to the authors

