## Supplementary material for "Lysosomal Storage Dysfunction, a Germline Variants Affecting Pancreatic Ductal Adenocarcinoma Development and Progression": LSD supplementary Figures and Figure legends 2023 0211 for Biorxiv.pdf

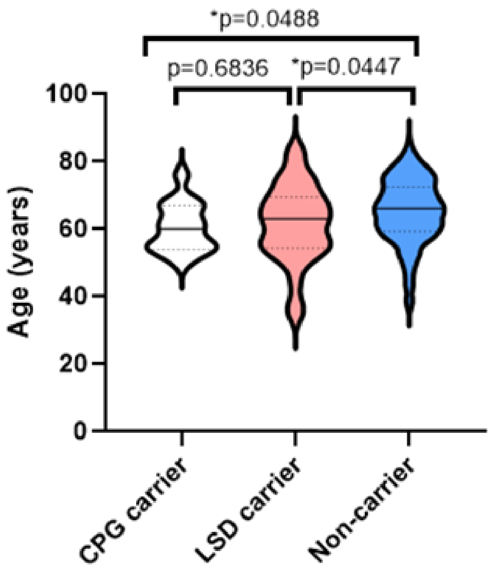


**Figure S1. Onset of age in PDAC patients with CPG or LSD carriers**

Age at diagnosis was compared in PDAC patients with CPG carriers, LSD carriers and non-carriers. PDAC, pancreatic ductal adenocarcinoma; CPG, cancer predisposition gene, LSD, lysosomal storage disease. p*<0.05.

A

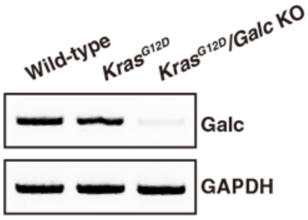


B

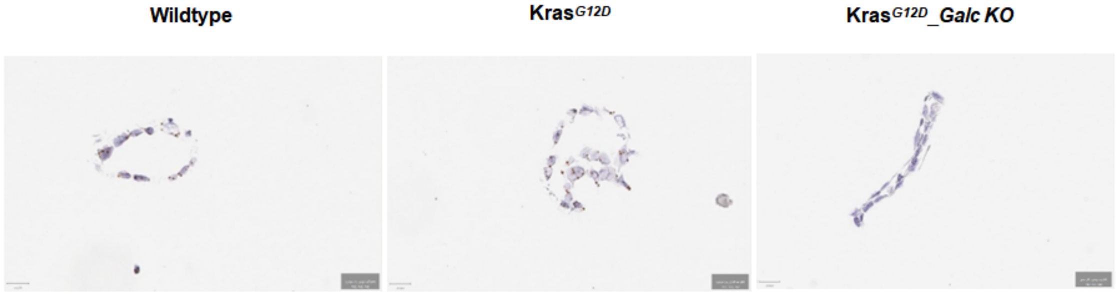


C

| | Positive cells (%) | Negative cells (%) | Result |
|---|---|---|---|
| Wildtype | 41 | 59 | Positive |
| Kras$^{G12D}$ | 42 | 58 | Positive |
| Kras$^{G12D}$/Galc KO | 3 | 97 | Negative |

**Figure S2. Evaluation of *Kras$^{G12D}$/Galc* knockout mouse pancreatic organoids**

(A) The expression of GALC in pancreatic organoids from wild-type, *Kras$^{G12D}$* knockout, *Kras$^{G12D}$/Galc* knockout mouses was examines by immunoblotting. GAPDH was used as an internal control.

(B and C) Mouse pancreatic organoids were immunohistochemically stained with GALC antibody, and the number of GALC-positive cells was counted.

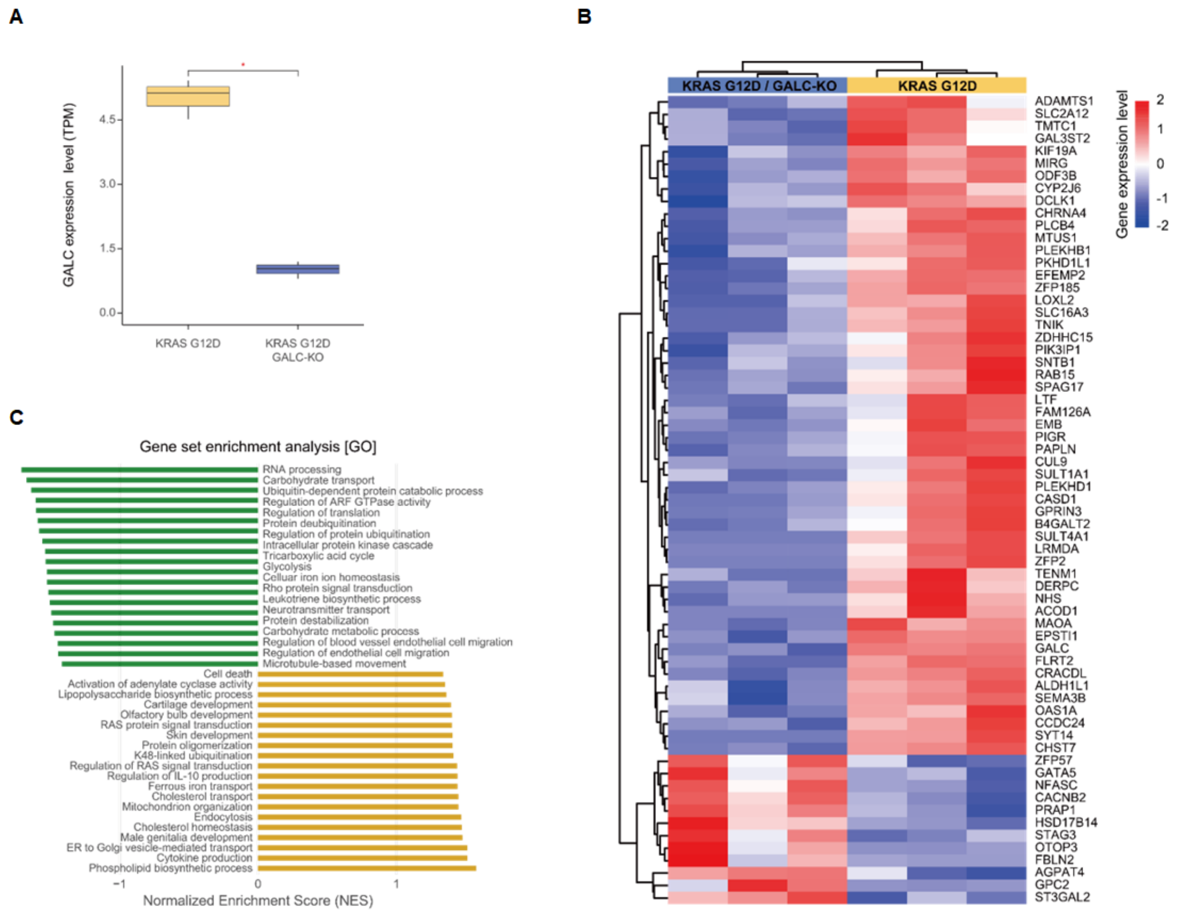


**Figure S3. Gene expression of KrasG12D/Galc knockout mouse pancreatic organoids**

(A) RNA sequencing was performed with pancreatic organoids from wild-type, *Kras*$^{G12D}$ knockout, *Kras*$^{G12D}$*/Galc* knockout mouses, and the expression level of GALC was accessed.

(B) Heatmap with RNA sequencing showed differentially expressed genes between *Kras*$^{G12D}$ knockout mouse and *Kras*$^{G12D}$*/Galc* knockout mouse.

(C) The biological process, cellular components and molecular function were identified from KEGG enrichment analysis. KEGG, Kyoto Encyclopedia of Genes and Genomes.

A

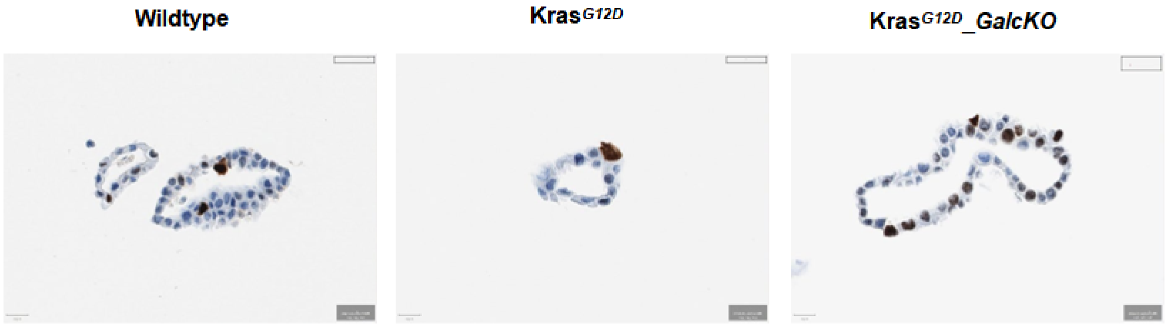


B

| | Positive cells (%) | Negative cells (%) | Ki67 index |
|---|---|---|---|
| Wild-type | 12 | 88 | 12 |
| $Kras^{G12D}$ | 9 | 91 | 9 |
| $Kras^{G12D}$/Galc KO | 32 | 68 | 32 |

**Figure S4. Increased proliferation in KrasG12D/Galc knockout mouse pancreatic organoids**

(A and B) In situ hybridization for GALC were performed with mouse pancreatic organoids and the percentage of Ki67-positive cells was calculated in wild-type, *Kras*$^{G12D}$ knockout, *Kras*$^{G12D}$*/Galc* knockout mouses.

A

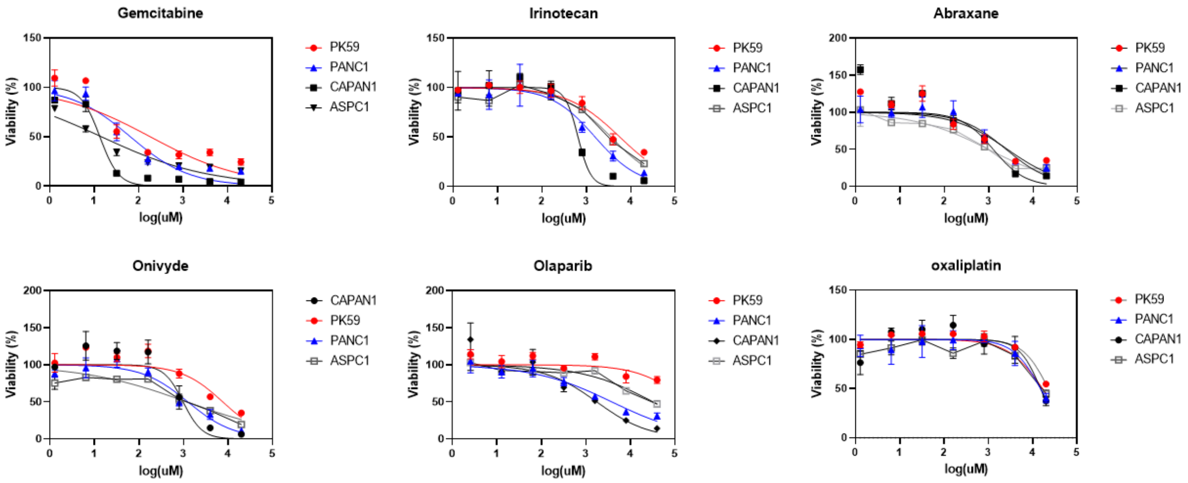


B

| IC50 (nM) | PK59 | PANC1 | CAPAN-1 | ASPC1 |
|---|---|---|---|---|
| Gemcitabine | 181.1 | 64.77 | 13.32 | 14.4 |
| Irinotecan | 5668 | 1574 | 649.6 | 3600 |
| Abraxane | 2645 | 2472 | 1263 | 984.1 |
| Onivyde | 7662 | 1256 | 980.8 | 1192 |
| Olaparib | 189813 | 3839 | 1758 | 32614 |
| Oxaliplatin | 22584 | 15020 | 15057 | 16816 |

**Figure S5. Responsed to drugs in human PDAC cell lines**

(A) PK59, PANC1, CAPAN1 and ASPC1 cells were treated with therapeutic drugs for 7 days, and cell viability was accessed using an adenosine triphosphate monitoring (triplicates).

(B) The half-maximal inhibitory concentration (IC50) values (nM) were calculated using GraphPad 8.0.
