## Supplementary material for "Lysosomal Storage Dysfunction, a Germline Variants Affecting Pancreatic Ductal Adenocarcinoma Development and Progression": LSD supplementary Table 1 2023 0211.pdf

### Supplementary Table 1. I

| Chromosome | Position |
|---|---|
| 22 | 51065136 |
| 22 | 51065757 |
| 17 | 78082617 |
| 17 | 78086479 |
| 17 | 78078931 |
| 17 | 78079575 |
| 14 | 88454867 |
| 14 | 88401093 |
| 14 | 88406259 |
| 3 | 33058311 |
| 12 | 102154949 |
| 12 | 102164207 |
| 12 | 102224453 |
| 16 | 1412062 |
| 7 | 65439577 |
| 15 | 72640414 |
| 5 | 74014629 |
| 8 | 43028883 |
| 3 | 50339902 |
| 3 | 50339936 |
| 3 | 50340307 |
| 4 | 995950 |
| 19 | 12760828 |
| 19 | 12766602 |
| 19 | 12774571 |
| 19 | 12777513 |
| 19 | 7592404 |
| 17 | 40693077 |
| 17 | 40695080 |
| 18 | 21141366 |
| 10 | 73579379 |
| 10 | 73585650 |
| 17 | 78194043 |
| 17 | 78187645 |
| 11 | 6415800 |
| 3 | 4458826 |
| 11 | 6637988 |

**Pathogenic variants identified in the Korean PDAC and Normal control**

| Reference Sequence |
|---|
| C |
| C |
| T |
| C |
| G |
| G |
| C |
| C |
| A |
| G |
| G |
| G |
| T |
| CCAA |
| G |
| G |
| C |
| C |
| C |
| G |
| GT |
| G |
| CAATGGGATGGCAAGGTTGTGAGCCTTGGATAAACCCCTCTGCCCTTGCTTCCACACCCCTCTCCCAGCCTGTC |
| G |
| G |
| CAT |
| A |
| G |
| G |
| CCTTATTGA |
| GAGGGCAGCGGGCTCAACGCTGGCAGGGCCCTCCCAGACCCAAGAGGGGCACCATCCTCTCCCGCACCACACC |
| A |
| TC |
| C |
| A |
| G |
| GTT |

| Alternative Sequence | Gene Symbol | gnomAD_AF | gnomAD_AF_EAS | SNP ID |
|---|---|---|---|---|
| T | ARSA | 1.20E-05 | 5.44E-05 | rs199476366 |
| T | ARSA | 1.53E-05 | 6.80E-05 | rs74315455 |
| A | GAA | 2.84E-05 | 4.00E-04 | rs747610090 |
| G | GAA | . | . | . |
| T | GAA | . | . | . |
| T | GAA | . | . | . |
| T | GALC | . | . | . |
| T | GALC | 2.00E-04 | 5.00E-04 | rs200607029 |
| G | GALC | 7.00E-04 | 8.50E-03 | rs138577661 |
| A | GLB1 | 1.60E-05 | 5.56E-05 | rs72555359 |
| A | GNPTAB | 3.98E-06 | 5.44E-05 | rs281865009 |
| A | GNPTAB | 1.19E-05 | 1.00E-04 | rs200646278 |
| A | GNPTAB | 4.13E-06 | 5.58E-05 | . |
| C | GNPTG | 9.29E-05 | 0 | rs193302849 |
| GC | GUSB | . | . | . |
| A | HEXA | . | . | . |
| T | HEXB | 6.00E-04 | 1.00E-03 | rs28942073 |
| T | HGSNAT | 2.84E-05 | 2.00E-04 | rs121908282 |
| T | HYAL1 | . | . | . |
| C | HYAL1 | . | . | . |
| G | HYAL1 | 4.03E-06 | 5.44E-05 | rs781974681 |
| A | IDUA | . | . | rs794727840 |
| G | MAN2B1 | . | . | . |
| C | MAN2B1 | . | . | . |
| A | MAN2B1 | . | . | . |
| C | MAN2B1 | 6.85E-06 | 9.37E-05 | . |
| C | MCOLN1 | . |  | . |
| A | NAGLU | 1.19E-05 | 0 | . |
| A | NAGLU | . |  | . |
| C | NPC1 | . | . | . |
| A | PSAP | . | . | . |
| ATATGT | PSAP | . | . | . |
| T | SGSH | . | . | . |
| T | SGSH | 1.69E-05 | 2.00E-04 | rs753472891 |
| [illegible]GGCCCAGAGCCTG[illegible] | SMPD1 | . | . | . |
| A | SUMF1 | . | . | . |
| G | TPP1 | . | . | . |

| Ensembl Transcript ID | Clinvar |
|---|---|
| ENST00000216124 | Pathogenic/Likely_pathogenic |
| ENST00000216124 | Pathogenic/Likely_pathogenic |
| ENST00000302262 | Likely_pathogenic |
| ENST00000302262 | Likely_pathogenic |
| ENST00000302262 | Likely_pathogenic |
| ENST00000302262 | . |
| ENST00000261304 | Likely_pathogenic |
| ENST00000261304 | Likely_pathogenic |
| ENST00000261304 | Pathogenic/Likely_pathogenic |
| ENST00000307363 | Pathogenic |
| ENST00000299314 | Pathogenic |
| ENST00000299314 | Pathogenic |
| ENST00000299314 | . |
| ENST00000204679 | Likely_pathogenic |
| ENST00000304895 | . |
| ENST00000268097 | . |
| ENST00000261416 | Pathogenic/Likely_pathogenic |
| ENST00000379644 | Likely_pathogenic |
| ENST00000266031 | . |
| ENST00000266031 | . |
| ENST00000266031 | . |
| ENST00000247933 | Pathogenic |
| ENST00000456935 | . |
| ENST00000456935 | . |
| ENST00000456935 | . |
| ENST00000456935 | Uncertain_significance |
| ENST00000264079 | . |
| ENST00000225927 | Likely_pathogenic |
| ENST00000225927 | . |
| ENST00000269228 | . |
| ENST00000394936 | . |
| ENST00000394936 | . |
| ENST00000326317 | . |
| ENST00000326317 | Likely_pathogenic |
| ENST00000342245 | . |
| ENST00000272902 | . |
| ENST00000299427 | . |

| **Clinvar_ref** |
|---|
| Metachromatic_leukodystrophy\|not_provided |
| Glycogen_storage_disease\|Glycogen_storage_disease,_type_II |
| Glycogen_storage_disease,_type_II |
| Glycogen_storage_disease,_type_II |
| . |
| Galactosylceramide_beta-galactosidase_deficiency |
| Galactosylceramide_beta-galactosidase_deficiency |
| PS-IV-B\|Infantile_GM1_gangliosidosis\|GM1_gangliosidosis_type_2\|C |
| Mucolipidosis\|Pseudo-Hurler_polydystrophy\|I_cell_disease |
| Mucolipidosis\|Pseudo-Hurler_polydystrophy\|I_cell_disease |
| . |
| . |
| . |
| sease\|Sandhoff_disease,_adult_type\|Sandhoff_disease,_juvenile_type\|r |
| trophy\|Mucopolysaccharidosis,_MPS-III-C\|Retinitis_pigmentosa_73\|n |
| . |
| . |
| . |
| . |
| . |
| Deficiency_of_alpha-mannosidase |
| Mucopolysaccharidosis,_MPS-III-B |
| . |
| . |
| . |
| . |
| Mucopolysaccharidosis,_MPS-III-A |
| . |
| . |
| . |

| Function | Protein change |
|---|---|
| missense_variant | p.Arg246His |
| missense_variant | p.Gly101Asp |
| missense_variant | p.Met439Lys |
| missense_variant | p.Ser619Arg |
| synonymous_variant | p.Thr182Thr |
| stop_gained | p.Glu192X |
| missense_variant | p.Ala66Thr |
| missense_variant | p.Val681Met |
| missense_variant | p.Leu634Ser |
| stop_gained | p.Arg457Ter |
| stop_gained | p.Arg1031Ter |
| stop_gained | p.Arg364Ter |
| start_lost | p.Met1? |
| inframe_deletion | p.Asn116del |
| frameshift_variant | p.Gln248fs |
| stop_gained | p.Gln350X |
| missense_variant | p.Pro417Leu |
| missense_variant | p.Pro283Leu |
| stop_gained | p.Trp162X |
| stop_gained | p.Ser151X |
| frameshift_variant | p.Leu27fs |
| splice_donor_variant | p.Pro192Leu |
| splice_acceptor_variant;splice_donor_variant | . |
| stop_gained | p.Ser579Ter |
| stop_gained | p.Gln237X |
| frameshift_variant;start_lost | . |
| splice_acceptor_variant | . |
| missense_variant | p.Gly292Arg |
| stop_gained | . |
| frameshift_variant | p.Phe194fs |
| splice_acceptor_variant;splice_donor_variant | . |
| frameshift_variant | p.Cys241fs |
| frameshift_variant | p.Arg23fs |
| missense_variant | p.Asp235Asn |
| frameshift_variant | p.Glu620fs |
| stop_gained | p.Gln276Ter |
| frameshift_variant | p.Gln263fs |
