## Supplementary material for "Lysosomal Storage Dysfunction, a Germline Variants Affecting Pancreatic Ductal Adenocarcinoma Development and Progression": LSD supplementary Table 2 2023 0211.pdf

**Supplementary Table 2. 42 lysosomal storage disorder associated genes**

| Gene Symbol | Ensembl ID | Chromosome | PDAC (%) | Normal (%) |
|---|---|---|---|---|
| *AGA* | ENSG00000038002 | 4 | - | - |
| *ARSA* | ENSG00000100299 | 22 | 0.2 | 0.1 |
| *ARSB* | ENSG00000113273 | 5 | - | - |
| *ASAH1* | ENSG00000104763 | 8 | - | - |
| *CLN3* | ENSG00000188603 | 16 | - | - |
| *CTNS* | ENSG00000040531 | 17 | - | - |
| *CTSA* | ENSG00000064601 | 20 | - | - |
| *CTSK* | ENSG00000143387 | 1 | - | - |
| *FUCA1* | ENSG00000179163 | 1 | - | - |
| *GAA* | ENSG00000171298 | 17 | 0.5 | 0.4 |
| *GALC* | ENSG00000054983 | 14 | 3.6 | 1.5 |
| *GALNS* | ENSG00000141012 | 16 | - | - |
| *GBA* | ENSG00000177628 | 1 | - | - |
| *GLA* | ENSG00000102393 | x | - | - |
| *GLB1* | ENSG00000170266 | 3 | - | 0.1 |
| *GM2A* | ENSG00000196743 | 5 | - | - |
| *GNPTAB* | ENSG00000111670 | 12 | - | 0.4 |
| *GNPTG* | ENSG00000090581 | 16 | 0.2 | - |
| *GNS* | ENSG00000135677 | 12 | - | - |
| *GUSB* | ENSG00000169919 | 7 | 0.5 | - |
| *HEXA* | ENSG00000213614 | 15 | 0.2 | - |
| *HEXB* | ENSG00000049860 | 5 | 1.2 | 0.7 |
| *HGSNAT* | ENSG00000165102 | 8 | - | 0.1 |
| *HYAL1* | ENSG00000114378 | 3 | 0.2 | 0.2 |
| *IDS* | ENSG00000010404 | x | - | - |
| *IDUA* | ENSG00000127415 | 4 | 0.2 | - |
| *LAMP2* | ENSG00000005893 | x | - | - |
| *LIPA* | ENSG00000107798 | 10 | - | - |
| *MAN2B1* | ENSG00000104774 | 19 | 0.2 | 0.4 |
| *MANBA* | ENSG00000109323 | 4 | - | - |
| *MCOLN1* | ENSG00000090674 | 19 | 0.2 | - |
| *NAGA* | ENSG00000198951 | 22 | - | - |
| *NAGLU* | ENSG00000108784 | 17 | 0.5 | - |
| *NEU1* | ENSG00000204386 | 6 | - | - |
| *NPC1* | ENSG00000141458 | 18 | 0.2 | - |
| *NPC2* | ENSG00000119655 | 14 | - | - |
| *PPT1* | ENSG00000131238 | 1 | - | - |
| *PSAP* | ENSG00000197746 | 10 | 0.2 | - |
| *SGSH* | ENSG00000181523 | 17 | 0.5 | 0.1 |
| *SMPD1* | ENSG00000166311 | 11 | - | 0.1 |
| *SUMF1* | ENSG00000144455 | 3 | - | 0.1 |
| *TPP1* | ENSG00000166340 | 11 | - | 0.1 |
