## Supplementary material for "Lysosomal Storage Dysfunction, a Germline Variants Affecting Pancreatic Ductal Adenocarcinoma Development and Progression": LSD supplementary Table 3 2023 0211.pdf

### Supplementary Table 3. List of antibodies used in the study

| Antibody name | Company (Catalog#) | Dilution |
|---|---|---|
| Anti-ki67 | Abcam (ab15580) | 1:200 |
| Anti-LAMP1 | Santa Cruz (sc-1992) | 1:200 |
| Anti-mTOR | Cell Signaling (#2972) | 1:1000 |
| Anti-phospho-mTOR | Cell Signaling (#2974) | 1:1000 |
| Anti-LC3B | Cell Siganling (#2775) | 1:1000 |
| Anti-Ubiquitin | Cell Signaling (#3933) | 1:1000 |
| Anti- SQTM1/p62 | Abcam (ab56416) | 1:1000 |
| Anti-actin | Abcam (ab8226) | 1:1000 |
| Fluor 647 conjugated Goat anti-Rabbit | Abcam (ab150083) | 1:200 |
| FITC conjugated Rabbit anti-Rat | Abcam (ab6730) | 1:200 |
